# Evaluating the effectiveness of facilitator-led mental health education program interventions to improve adolescent mental wellbeing

**DOI:** 10.1101/2025.02.16.25322348

**Authors:** Evan Chang, Ruslan Bayliyeva, Yiliang Zhu

## Abstract

**Background:** In the last decade, youth mental wellbeing has plummeted across the nation as 1 in 5 high schoolers have seriously considered suicide, while the suicide rate increased by nearly 40%. Especially in the wake of COVID-19, a loss of independence and increased social isolation contributed to 70% of young people reporting worsening mental health coupled with a significant rise in hospitalization due to self-injury. Tele-mentoring and train the trainer approaches to train school mental health educators are increasingly popular interventions to target preventive wellbeing education to combat conditions such as anxiety, depression, emotional regulation and stress. In particular, the Project Evaluation of Community Healthcare Outcomes (ECHO) uses an evidence-based model to support providing mental health services in its hub-and-spoke model using didactic and case-based module learning.

**Objective:** The purpose of this systematic review is to evaluate the effectiveness of mental health educational interventions led by providers/educators, which use tele-mentoring and other community learning models (e.g., the ECHO model). Furthermore, we evaluated whether there is improvement in mental wellbeing and/or an increase in subject knowledge, self-efficacy, satisfaction, community building, and acceptability.

**Methods:** A systematic review was performed based on PRISMA (Preferred Reporting Items for Systematic Reviews and Meta-Analyses) and PROSPERO (International Prospective Register of Systematic REVIEW) guidelines. We searched for key words using PubMed, Embase, Web of Science, Ovid, and PsycINFO in September 2024. Interventions were included if the study was published from 2000 until the present, conducted within the United States, reached a population of school or non-school-based educators teaching adolescents ages 10-18, and reported on general mental wellbeing (i.e., anxiety, depression, suicidal ideation, stress, mindfulness, and emotional regulation).

**Results:** After retrieving a total of 2603 articles from database searching, further screening reduced the number of articles that met the inclusion criteria to 12. All articles used a train the trainer approach where facilitators educated a teacher population; 6 articles went further to examine outcomes from teachers educating an adolescent population.

Among the articles which only educated a teacher population, program satisfaction, fidelity, engagement, and curriculum literacy reached highs of 100%, 83.3%, 100%, and 66.7%, respectively. Qualitative responses show that adolescents who participated had positive self-reported effects on social resilience and emotional regulation through a greater understanding of dialectical behavior therapy. These findings suggest that teachers found the training applicable to their own teaching environments, and the material both adaptable and conducive to fostering a positive and emotionally healthy classroom environment. However, measured impact among adolescents still remains unclear without direct outcomes responded by the adolescents themselves. In the teacher and adolescent training programs, there were more revealing with specific outcomes better tailored to the program and response for adolescents. Among the articles which also educated an adolescent population,100% reported high curriculum program knowledge retention rate in literacy for either their adult educator or adolescent cohorts. Therefore, these varying and nuanced findings show the need for further research in individual engagement and pre-existing risk factors in determining program success.

**Conclusions:** Interventions enhancing school mental health professional or general school educator mental health literacy through an ECHO-like model could potentially lay the groundwork for improved youth wellbeing and engagement, underscoring the importance in fostering resilience among community wide evidence-based practices. These interventions demonstrated most notably improved satisfaction and curriculum understanding levels as compared to their control group baseline scores, although the success of which may be limited to the number of studies implemented, the role of student engagement, and the mode of measuring outcomes.

## Introduction

In the United States, mental health problems even before the COVID-19 pandemic, have been the leading cause of poor life outcomes in young people as one in five children aged 3–17 reported having a mental, emotional, developmental, or behavioral disorder.^1^ In 2019, a survey by the Centers for Disease Control found that more than one in three high school students experienced persistent feelings of sadness or hopelessness that year—a 40% increase from the numbers recorded a decade before.^1^ Furthermore, one in six youth reported making a suicide plan during that year—a 44% increase over 2009.^1^ And after the start of the pandemic, in 2021, the American Academy of Pediatrics, American Academy of Child and Adolescent Psychiatry, and Children’s Hospital Association declared a national emergency in child and adolescent mental health^2^. Over 75% of young people in primary services and 80% in the general population reported that the pandemic negatively affected their work, non-work life, and general mental health. These statistics are consistent with COVID-19 youth surveys conducted globally.^3^According to the World Health Organization, globally, about one in seven adolescents have a mental health disorder, depression, or anxiety. Behavioral disorders are among the leading causes of illness and disability among 10- to 19-year-olds,^3^ while suicide is the fourth leading cause of death in adolescents worldwide.^3^

Unfortunately, surveys have shown that 70–80% of adolescents with mental illnesses do not receive the mental health services they need.^4,5^ The main reason is the delay of an early intervention caused by a lack of full-time and part-time teachers for mental health service resources in schools^6^, low levels of specialization, limited mental health service resources, and low mental health literacy in students.^5 7^Among the available school staff, many are increasingly seeking to implement evidence-based school mental health services to promote student mental health, yet accessing programming and support can be quite difficult.^8^ Even strategies to avoid such challenges, like one-time professional development, often fail to be effective or sustainable. In response to challenges associated with online learning, novel strategies are useful for expanding access to information about evidence-based school mental health care, specifically tele-mentoring models. Tele-mentoring is a form of training and support practice that is held through virtual sessions that emphasize collaborative relationships between content experts and participants alike. Tele-mentoring training programs that use sequential online learning modules combined with interprofessional tele-mentoring, e.g., the Project Extension for Community Healthcare Outcomes (ECHO) model, have become popular in influencing provision of school mental health services.^9^ Developed to increase physicians’ knowledge about best practice instruction through didactic and case-based virtual mentoring, the structure of the Project ECHO model involves regularly scheduled, live, virtual meetings where content experts (typically four or five, referred to as the *hub team*) and healthcare providers or educators (typically 20–30, referred to as the *spokes*). Each meeting consists of short introductions (approximately 5 min); a presentation about the topic related to evidence-based care given by a hub team member (approximately 20 min); and a case presentation from one of the spokes. During the case presentation, a facilitator invites the participants to ask clarifying questions along with specific recommendations (approximately 30 min). Studies and reports of the Project ECHO model frequently show moderate positive changes in participants’ self-efficacy and knowledge about evidence-based practices as well as patient outcomes.^10,11^

The purpose of this systematic review is to evaluate the effectiveness of mental health educational interventions led by providers/educators, which use tele-mentoring and other community learning models (e.g., the Project ECHO model). Furthermore, we evaluated whether there is improvement in mental wellbeing and/or an increase in subject knowledge, self-efficacy, satisfaction, community building, and acceptability.

## Methods

### Search strategy and Article selection

Following PRISMA (Preferred Reporting Items for Systematic Reviews and Meta-Analyses) and PROSPERO (International Prospective Register of Systematic REVIEW) guidelines, we searched for key words using PubMed, Embase, Web of Science, Ovid, and PsycINFO in September 2024. Our initial search criteria were included if the study was an original research article, published from 2000 until the present, provided the full-text in English, conducted within the United States, reached either a population of school-based educators/counselors that simply participated in the training session labeled Group 1 studies (Table 1) or a population of teachers executed the trainings to adolescents ages 10-18 labeled Group 2 studies (Table 2), performed without conducting a pre-diagnosis test (such as determining if participants have any learning disabilities are pre-existing anxious qualities), did not conduct a follow-up study, and reported on general mental wellbeing (i.e., anxiety, depression, suicidal ideation, stress, mindfulness, and emotional regulation).

**Table 1.** Group 1 articles with teacher intervention only (See references ^12,13,8,9,14,15^)

| Author, Year, Title | Curriculum | Study Design | Intervention details | Outcomes evaluated | Results |
| --- | --- | --- | --- | --- | --- |
| Adrian, 2017, Enhanced "Train and Hope" for Scalable, Cost-Effective Professional Development in Youth Suicide Prevention | training and hope professional development model knowledge, attitudes, and behavior surrounding suicide assessment intervention developed by child psychologists and psychiatrists. Content included descriptive epidemiology, assessment strategies, management of suicide risk, and special populations and cultural considerations. Combination of didactic and active learning techniques, | n/a | 6 hour continuing education training for 83 Seattle Children's hospital mental health practitioners. Also using email reminder system | knowledge, attitudes, and behavior towards suicide assessment and management. and likelihood of practitioners in condition group to implement standard suicide assessment | All practitioners demonstrated increase in suicide assessment knowledge and attitudes for engaging in suicide risk assessments from pre- to posttest, and gains were maintained at the 3 month follow-up highly relevant, satisfied with effectiveness of the trainers, significant increase in suicide assessment knowledge, significant increase in attitudes. No differences found by the email reminder condition. |
| Hambrick, 2023, Disseminating early interventions for disaster mental health response using the ECHO model | ECHO used for disseminating early disaster interventions: Psychological First Aid and Skills for Psychological recovery to school professionals throughout rural, disaster-affect communities. PFA did tier 1 and SPR did tier 2. Used ECHO models teaching elements. | n/a | 164 participants for pre training webinar and 84 participants for four part training | participation, satisfaction, learning, competence, and performance | high participation, high satisfaction, high use at 1 month follow up, high competence/perceived learning, performance was hard to measure |
| Lyons, 2024, Applying the Extension for Community Healthcare Outcomes (ECHO) Model to Promote Collaborative and Effective School Mental Health Services | set of training activities (sequential online learning modules combined with interprofessional telementoring ECHO model) influenced provision of school mental health services. All participants had access to 12 online learning modules. 1st: evidence based and equitable practices. 2nd: effective and cultural responsive practices. 3rd: interprofessional collaboration. Topics included using OARS model, behavioral activation, racial trauma, and stress first aid. | mixed methods | nine 60 min ECHO sessions. 46 school counselors, nurses, psychologist, and social workers. First group included 29 and met monthly. Second group met twice monthly in September. Facilitated by research associated certified in school psychology. There were focus groups | perceptions of feasibility and appropriateness of the ECHO model (acceptability, relevance, and applicability), content knowledge, self-efficacy | high participation, statistically significant self efficacy and knowledge of evidence based practices. Positive reactions to experience and behavior change (way they will prompt change and provide services in schools). Sense of community and connection was built |
| Lyons, 2022, Supporting School Mental Health Providers: Evidence from a Short-Term Telementoring Model | two sets of online learning modules, 8 total on evidence based practices, multi-tiered systems of support, CBT, culturally responsive practices for supervising. Content was created by researchers and online instructional designers. | quasi experimental design | All 57 school staff access to 8 asynchronous, online modules. Sub group had access to monthly ECHO sessions that lasted for 60 minutes. Study was part of a larger ongoing state wide research practice partnership. Before the study, there were 4 monthly ECHO sessions that covered behavioral activation, motivational interviewing, and racial trauma. New cohort was found for the study obviously. | Individual: engagement, satisfaction, and knowledge | high staff engagement and satisfaction with the modules but no observed differences in knowledge gained |
| Taylor, 2022, Utilizing a Telementoring Model to Promote the Evidence-Based School Counseling Model |  | mixed method intervention | 42 school mental health professionals were given access to 12 online learning modules. Nine ECHO participants also participated in post-training focus groups. There were 9 ECHO session 60 minutes per session. 28 were in the ECHO and Modules condition and 16 were in the modules only condition. | participation, knowledge, participant's application of evidence based application, multicultural knowledge, interprofessional collaboration, and behavior/implementation | highly engaged, scored higher in all three measures of multicultural competence, and interprofessional application. ECHO participants had largest growth in application of EBP and multicultural competence. Qualitative data supports that there was greater identification of cultural and environmental factors, interprofessional collaboration, greater connection through ECHO community, decreased professional isolation, improvements in professional advocacy. Module + ECHO participants had greater improvements in collaborative processes and had increased knowledge EBP and cultural competent practices |
| Zhang, 2023, Connecting Behavioral Health Specialists With Schools: Adapting a Telementoring Series During Covid-19 | Telehealth Rural Outreach for the Children of Kansas partnered with ECHO, created the Function Friday for Better Behavior designed to facilitate understanding application of behavioral plans for school. | n/a | Eight sessions about functional behavior twice a month over a 4 month period. 1 hour ECHO sessions. 35 participated | Knowledge and skill development, access to specialist team, satisfaction of ECHO format, engagement, application of content | high improvement of knowledge or skills, high access to specialist team, satisfaction with ECHO format, high application of content. |

**Table 2.** Group 2: articles which also evaluated adolescent outcomes (See references^16,17,18,19,20,21^)

| Author, Year, Title | Curriculum | Study Design | Intervention details | Outcomes evaluated | Results |
| --- | --- | --- | --- | --- | --- |
| Dawes, 2019, Creating Supportive Contexts for Early Adolescents during the first year of middle school: impact of a developmentally responsive multi-component intervention | Teachers received training in the Behavioral, Academic, and Social Engagement (BASE) classroom management model. Looking at how middle schoolers can adjust to academic, behavioral, and social domains. | cluster randomized controlled trial | Teachers met for 9 hour training introductions and then completed 6 online modules. 12 middles schools, 122 6th grade teachers, and 1537 6th graders in the intervention were involved in the study. Length unknown | self-reported outcome on students' school belonging, peer protective ecology, defiance, social anxiety, emotional symptoms | Vulnerable subgroups and lower economic status reported had improved academic engagement and success and exhibited less defiance and bully protective ecology. Positive classroom environments were created for lower income students. School valuing and school belonging in the spring semester were significantly and positively related to perceptions of peer norms for effort and perceptions for peer protection. Results revealed only moderation effectson students' self-reported perceptions of the school context or adjustment indices. This suggests that intervention effects are nuanced in that there may be particular benefits to more vulnerable youth in metropolitan school settings such as ethnic minority students and students with lower economic status. |
| Frank, 2021, The Effectiveness of a Teacher-Delivered Mindfulness-Based Curriculum on Adolescent Social-Emotional and Executive Functioning | manualized mindfulness based program Learning to BREATHE (body, reflections, emotions, attention) on social-emotional wellbeing, mental health. Units cover somatic awareness/intro to mindfulness, using self-talk, explores how emotions affect thoughts, and understanding stress. | quasi-experimental control group trial | two intervention teachers had a 12 lesson sequence over a 6 week period with coaches who taught delivering the program. Coaches also made one classroom visit with each teacher. 251 11th grade students received material in 12 sessions, two sessions for each unit | mindfulness, emotional regulation, self-compassion, somatization, sleep, depression, anxiety, social connectedness, mind-wandering, growth mindset, rumination, stress, risk taking, working memory etc. | There were not intervention main effects on most of the adolescent self-report measures. In addition, the results of the moderation analyses suggest that beneficial program effects within the treatment group are influenced by the degree to which students report actively engaging in mindfulness practices outside of the classroom setting. Students who reported higher rates of practice (greater than once per month) showed significant small-to-moderate interaction effects on measures of emotional awareness, emotional clarity, impulse control difficulties, social connectedness, mind-wandering, substance use, stress reduction, and self-compassion. Unlike self-report measures, there was no significant interaction effects with practice for executive function outcomes. Although the present study did integrate mindfulness practice into the regular school day, findings suggest that adolescents' own choices and willingness to engage in practices outside of the regular school setting may be key to the success of universal school-based approaches. |
| Lindow, 2020, Feasibility and Acceptability of the Youth Aware of Mental Health (YAM) Intervention in US Adolescents | Youth Aware of Mental health (YAM) is a suicide prevention educational program that raises awareness about risk and protective factors such as knowledge about depression and anxiety as well as enhancing emotional resiliency | randomized control trial | YAM facilitators received a 5 day course. In 3-5 weeks, the facilitators instructed 5 hourly sessions for 1878 adolescents, three role plays and two interactive suicide prevention didactics | feasibility, acceptability, and fidelity of the YAM material : consent, participation, intervention adherence, completion by the give schedule, safety | This study found YAM feasible to implement in US schools. Students, parents, and school staff highly satisfied with the YAM program. The recruitment of schools was feasible in both states, with high proportions of schools agreeing to participate. The current study found that YAM was delivered with a high degree of fidelity to the program's content. Attendance of 86% available sessions in Montant was considerably higher than in the only 3 similar intervention studies... In this study, 93% of consented students participated in baseline surveys and 84% participated in both surveys, indicating low attrition rates. |
| Martinez, 2021, Effects of Dialectical Behavioral Therapy Skills Training for Emotional Problem Solving for Adolescents (DBT STEPS-A) Program of Rural Ninth-Grade Students | Staff Training for Emotional Problem Solving in Adolescents (STEPS-A): mindfulness and emotional regulation skills. It is structured into four modules: mindfulness, distress tolerance, emtoion regulation and interpersonal effectiveness. The training takes elements from dialectical behavior therapy. It encopasses 30 lessons, 50 minutes each in a environment for up tol 30 pupils. | quasi experimental control group design | 2, 6 hour training delivered to 8 school counselors. Over twelve weeks in 30 60 minute modules, these facilitators delivered the material to 94 9th grade students in a high school at 4 health/PE classes, 2 were control and 2 weren't. | social emotional outcomes (social resiliency, emotional regulation difficulties) along with dialectical behavior therapy skill understanding, acceptance, and self-efficacy | In addition to improvements in self-reported social resiliecn and reduction in regulation difficulties, DBT also increased emotional regulation and skill understanding (lower ratings of depression, anxiety and social stress and an increase in the ability to verbalize emotions). By learning how to teach and model DBT skill use, school counselor perspectives and practices may have shifted and also reinforced skill learning. |
| Schilling, 2015, The SOS suicide prevention program: Further evidence of efficacy and effectiveness | Signs of Suicide (SOS) program goals are to (1) to increase an understanding of depression as an illness and suicide as a behavior related to untreated/poorly managed depression, (2) to improve attitudes toward intervening with peers who are experiencing symptoms of depression and might be thinking about suicide, and (3) to encourage youth who are contemplating suicide to seek help | randomized control design | School counselors and social work staff received a 1 day training on the material. Over three months, they administered this to 9th graders in 16 high schools, 1052 completed both pre-and post-test | self-reported suicidal ideation, suicide planning, and suicide attempts, knowledge about anxiety and depression and suicide | Participation in the SOS program was associated with lower rates of suicide attempt at 3 months following the program but was not associated with changes in suicidal ideation. In addition, the current study extended previous research by measuring self-reports of suicide planning, but did not find a main effect of program participation on this outcome. The data suggest that the program may be more effective among high-risk students in terms of reducing the probability of suicide planning |
| Swartz, 2017, School-based curriculum to improve depression literacy among US secondary school students: a randomized effectiveness trial | The Adolescent Depression Awareness Program (ADAP) is a universal school-based depression education program to increase depression literacy as the first step in encouraging depressed youths to seek treatment. ADAP delivers the core message that depression and bipolar disorder are treatable medical illnesses and that concerned individuals should seek help. | matched pairs design | ADAP training is 6 hours long for teachers and school personnel which include 3681 intervention 9th graders from 66 high schools received 3 hour instruction broken up into 2-3 classroom periods. Topics include: identifying symptoms of depression, understanding the process of medical decision-making, seeing parallels between depression and other medical illnesses, recognizing suicide as a potential consequence of depression, and understanding that depression is a treatable medical illness. | depression literacy, mental health stigma, mental services receipt | ADAP resulted in significantly higher levels of depression literacy among participating students than did waitlist controls, after adjusting for pretest assessment depression literacy. Overall, ADAP did not significantly affect stigma. After ADAP, students approached 46% of teachers with concerns about themselves or others. Of students who reported the need for depression treatment, 44% received treatment within 4 months of ADAP implementation. |

For the identification of the Group 1 studies, we used multiple combinations of key terms to arrive at our final number of articles. The search query for the search that produced the greatest number of accepted articles for PubMed are as follows:

((adolescent [MeSH Terms]) OR (children [MeSH Terms]) OR (teenager [MeSH Terms)) AND ((mental health) OR (depression) OR (anxiety) OR (suicid*) OR (mental well-being) OR (mental wellbeing) OR (emotional regulation)) AND ((module*) OR (mental health/education[MeSH Terms]) OR (Curriculum*[MeSH Terms]) OR (tele-mentor*) OR (telementor*) OR (instruction*)) AND ((intervention*) OR (ECHO) OR (intervention study)) NOT ((systematic review) OR (meta-analysis) OR (literature review)) NOT ((website) OR (internet) OR (online) OR (app)) NOT ((parent) OR (university student) OR (adult) OR (autism) OR (eating) OR (pregnancy) OR (ADHD) OR (disorder*))

Group 1 search query for the other databases are as follows:

(((adolescent or children or teenager) and (mental health or depression or anxiety or suicid* or mental well-being or mental wellbeing or emotional regulation) and (module* or mental health education or Curriculum* or tele-mentor* or telementor* or instruction*) and (intervention* or ECHO or intervention study)) not (systematic review or meta-analysis or literature review) not (website or internet or online or app) not (parent or university student or adult or autism or eating or pregnancy or ADHD or disorder*))

For Group 2, the search query for the search that produced the greatest number of accepted articles for PubMed are as follows:

((Evidence-based practice [MeSH Terms]) OR (ECHO) OR (case-based) OR (school mental health services [MeSH Terms]) OR (capacity building) OR (mental health/education[MeSH Terms]) OR (implementation science [MeSH Terms])) AND ((Curriculum*[MeSH Terms]) OR (instruction*) OR (intervention*) OR (intervention study) OR (tele-mentor*) OR (telementor*) OR (tele-education)) AND ((suicide prevention [MeSH Terms]) OR (mental health) OR (depression) OR (anxiety) OR (suicid*) OR (mental well-being) OR (mental wellbeing) OR (emotional regulation)) AND ((school mental health providers) OR (school counselors) OR (counselors [MeSH Terms]) OR (teachers) OR (instructors) OR (educational personnel [MeSH Terms]) OR (health educators [MeSH Terms]) OR (school staff)) NOT (systematic review) NOT (meta-analysis) NOT (literature review)

Group 2 search query for the other databases are as follows:

(((Evidence-based practice) OR (ECHO) OR (case-based) OR (school mental health services) OR (capacity building) OR (mental health/education) OR (implementation science)) AND ((Curriculum*) OR (instruction*) OR (intervention*) OR (intervention study) OR (tele-mentor*) OR (telementor*) OR (tele-education)) AND ((suicide prevention) OR (mental health) OR (depression) OR (anxiety) OR (suicid*) OR (mental well-being) OR (mental wellbeing) OR (emotional regulation)) AND ((school mental health providers) OR (school counselors) OR (counselors) OR (teachers) OR (instructors) OR (educational personnel) OR (health educators) OR (school staff)))

Two reviewers (EC and RB) conducted independent dual review of identified references first by title and abstract, then by full text. At each stage, disagreements about inclusion or exclusion were adjudicated by a third investigator (YZ).

### Analysis

Articles included after full-text screening were divided and abstracted by database source and study type (Group 1 or Group 2) into a data abstraction file. We categorized the studies further by the outcomes that were evaluated (i.e. satisfaction, self-efficacy, feasibility) along with the study design.

### Results

After retrieving a total of 2603 articles from database searching, we narrowed the articles to 45 relevant articles after screening the full text and removing duplicates. Further screening reduced the number of articles that met the inclusion criteria to 12. All articles used a “train the trainer” approach where facilitators educated a teacher population; 6 articles went further to examine outcomes from teachers educating an adolescent population. (Figure 1)

**Figure 1.**
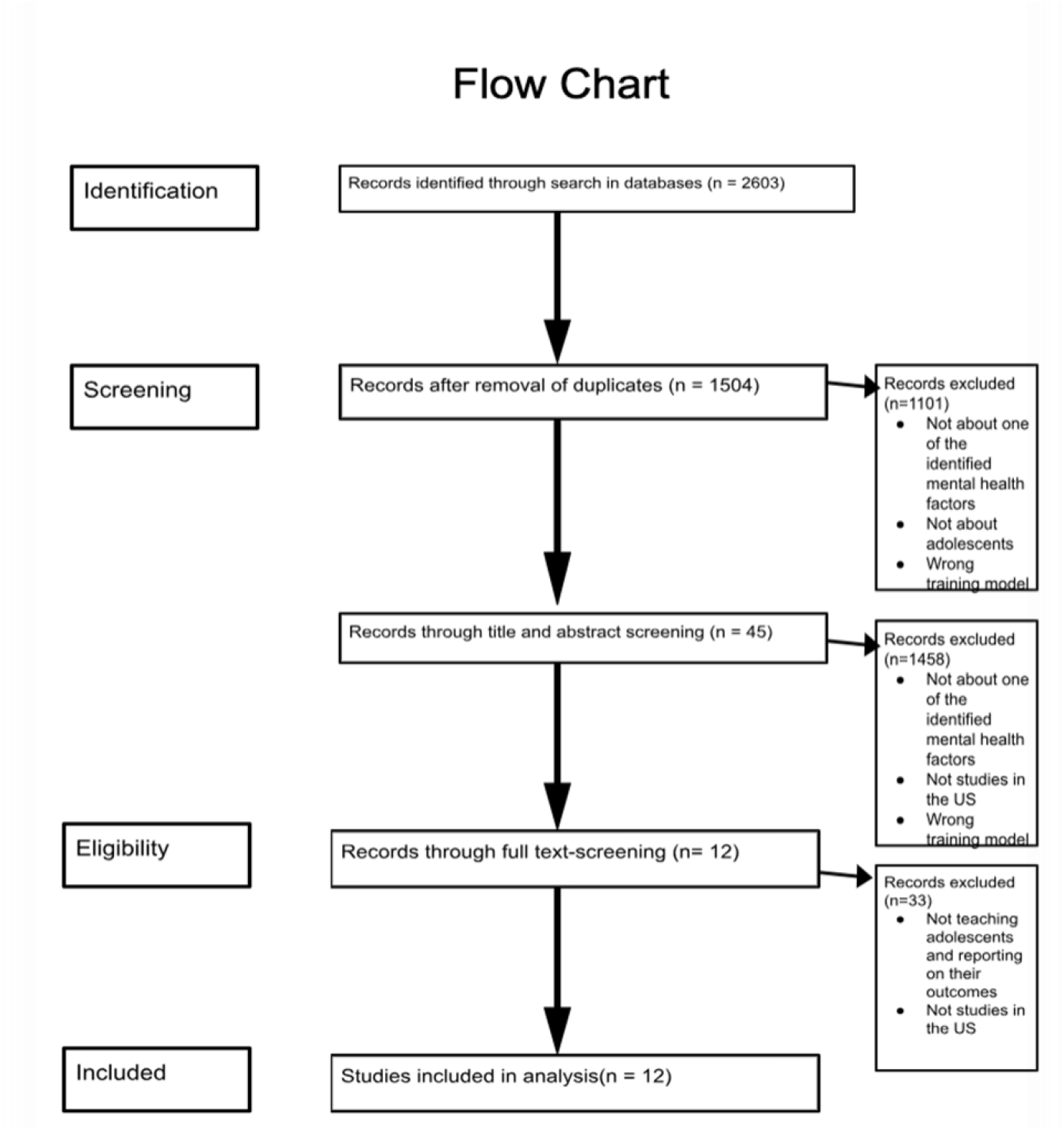
Flow chart of data abstraction process

### Satisfaction (n = 6)

All studies had a majority of participants that expressed moderate to high satisfaction with the given curriculum provided to them. This data was obtained through a combination of post-session, post-program, and 3-month follow-up questionnaires regarding their satisfaction and continued interest in not only the material discussed, but the structure and format of the modules. Some questions included: *Was there an adequate balance between didactic, case presentation, and interactive components of the sessions*? This category also included whether the studies’ participants were satisfied with the case presenters’ instructional methods and teaching formats.

### Fidelity/future implementation (n = 6)

Five out of six studies (83.3%) demonstrated an interest and willingness for future implementation of the program among their respective groups of adolescents. Sometimes referred to as positive behavior change, fidelity measures the enthusiasm to prompt change and act on the material, whether speaking to school administrators about the possibility of future instruction or independently planning to utilize the modules for immediate lessons. Furthermore, these participants found that the material they learned were moderately to highly applicable to their own lesson plans and can be easily integrated into their teaching style or school environment.

### Participation and engagement (n = 5)

Five out of six (83.3%) studies showed high participation and high engagement among participants in their sessions. Through qualitative observations from facilitators in meetings, session attendance reports by week, or well-developed case presentations, participants were contributing to the collaborative atmosphere crucial to a positive learning environment. Specifically in Taylor et. al.^14^ this increased engagement directly resulted in greater interprofessional collaboration, greater connection through the Project ECHO community, decreased professional isolation, and improved scores in competency and curriculum knowledge.

### Curriculum literacy (n = 6)

Four out of six (66.7%) studies showed moderate to high retention of curriculum otherwise known as curriculum literacy among participants. As the design for Group 1 studies are for the participants to replicate the instruction of similar or the same module among their own learning groups; therefore, curriculum literacy is one of the most crucial learning outcomes from these studies. Through both Likert-scale self-reporting and post-program/post-session quizzes, participants scored moderately high to high regarding the content which they were taught.

### Curriculum literacy (n = 6)

Six out of six studies’ adolescent participants demonstrated moderately high to high curriculum literacy after the program’s end. Differing from the Group 1, curriculum literacy outcome measure reporting, Group 2 participants not only received their instruction from only partial experts in the field (the newly trained participants) but the only measure for their literacy were through quizzes and tests, often considered a more accurate reading compared to simple self-reporting. Therefore, this statistic helps to prove the effectiveness of this “train-the-trainer” approach through a more objective means.

### Independent outcomes

Outside of curriculum literacy, Group 2 studies all measured separate, more specific outcomes that could not be standardized and grouped together like Group 1 outcomes because of their disparate and unrelating nature. (The only outcome reported in more than 1 study was acceptance.)

(P = positive effect)

(N = neutral effect)

- School belonging/social connectedness - P
- Peer protection - P
- Defiance - P
- Anxiety - P
- Working memory – P
- Stress - N
- Risk taking - N
- Growth mindset - N
- Sleep - N
- Executive functions - N
- Social resiliency - P
- Acceptance (2) - P
- Self-efficacy - P
- Suicidal ideation/planning - P
- Stigma - N

In the adolescent populations, there were improvements in decreasing social anxiety (especially among girls), creating a positive classroom environment, establishing a sense of school belongingness that protected students against bullying and peer pressure (peer protection), increased social resilience along with emotional regulation and shifted perspectives. In particular, the Dawes et. al.^21^ and Lindow et. al.^17^ studies found positive results from both their adolescent and teacher participants; Dawes et. al.^21^ most importantly observed a correlation between significant improved in outcomes among vulnerable subgroups and lower economic status groups while Lindow et. at.^17^ received feedback from a highly satisfied, engaged, and emotionally literate set of participants. Ethnic minority students at the end of the program exhibited less defiance and incorporated a more positive mindset to classroom collaboration and reflective nature to their inner emotions. Even though their reports from more middle-class youth from other school environments can be more nuanced and less revealing, Dawes and their team brought to light particular benefits attractive and effective for this subset of students.^21^Furthermore, Lindow et. al. found high participation rates within their sessions— 86% of participants attended all of the sessions and 84% of students completed both the baseline and post-program survey.^17^ Staff, too, were highly satisfied with the program as seen through the high proportion of fidelity for the program’s content.^17^

However, other study findings were not as conclusive as seen through Frank et. al.^16^and Schilling et. al.^19^ Contrary to expectations, Frank et. al. found little effects on the adolescent self-report measures; even though there were instances of improved selective attention, executive functions, and emotional moderation among certain individuals, there were little signification interaction effects among the whole of the sample across the program.^16^ The success of the program depended heavily on the rates of practice (greater than once per month attendance), concluding that adolescents’ own choices and engagement in the practices will dictate these universal school-based approaches more than the integration of mindfulness practices will. Similarly in Schilling’s study, the study did not have an effect on their main measurable outcome, suicidal ideation.^19^ Program participation, curriculum literacy, or engagement did not have an effect on self-reported suicide planning, suggesting that the program may be only effective among high-risk students in reducing the probability of suicide planning compared to the general population.^19^

## Discussion

Our systematic review identified 12 articles that evaluated the effectiveness of facilitator-led mental health programs to improve adolescent wellbeing that were divided into two groups, Group 1 and Group 2, because of their notable differences between the two implementation approaches. Group 1, where the program solely taught teachers without the intention to receive adolescent feedback and participation found high satisfaction, engagement, and curriculum literacy among the teacher participants. Through this approach, the findings suggest that teachers found the training applicable to their own teaching environments, and the material both adaptable and conducive to fostering a positive and emotionally healthy classroom environment. Through high scores in satisfaction, fidelity, engagement, and curriculum literacy, teachers have shown their acceptance for a professional training program to enable greater collaboration and confidence among educators.

As we know, Group 1 is a simpler and less complex group of studies as they only involve teachers who naturally are more inclined to engage and participate in such studies, introducing elements of bias. The outcomes will always be subjective as the effectiveness of such programs ultimately depend on teachers’ translation of their learning to the adolescent population. Therefore, without more objective adolescent outcome measures in these studies, we are limited in scope to truly see the effect of a comprehensive evaluation on student mental wellbeing.

In Group 2 studies, where trained teachers subsequently instructed adolescents, we can see more direct evidence of adolescent benefits; and despite the complications in this approach, the Group 2 studies still reported relatively high levels of curriculum literacy, despite teachers not being experts in mental health and students taking this program as a graded required class in school. This suggests that facilitator-led instruction can still be successful at transferring knowledge from teacher to adolescent. Additionally, Group 2 studies measured a diverse array of adolescent specific outcomes such as social resilience, working memory, growth mindset, and peer protection. Although not every outcome was considered of significant positive effect, the breadth of such specific outcomes that can only be answered from adolescent participants is the first big step in this process. Studies such as Dawes et. al. found particularly strong outcomes for vulnerable subgroups and the potential for programs to target minority and low economic status strata of youth.^21^

Despite high curriculum literacy, such specific outcomes were not as effective: Frank et. al. and Schilling et. al. are examples of how studies are not always universal in improving mental wellbeing as program impact are often directly rooted in student engagement and baseline risk factors.^16,19^ Frank et. al. found that self-directed engagement plays a crucial role in improving efficacy of programs^16^; Schilling et. al. discovered no significant impact on suicidal ideation, suggesting that only high-risk adolescents can be affected by program material^19^. Therefore, these varying and nuanced findings show the need for further research in individual engagement and pre-existing risk factors in determining program success.

Our review has some limitations. Although we used multiple large databases of indexed references on the biomedical topics, with any systematic literature review, our search strategy may have missed some relevant articles. For the sake of feasibility and remaining unbiased, we limited the geographic scope to only articles in the United States; in attempting to remove cultural differences, we also not only shrunk the sample size, but potentially missed key articles with helpful insights and supplementary findings. For example, there is a general lack of all facilitator-led mental health programs across the nation, specifically the “train-the-trainer” Group 2 studies. Yet, even with our limited small sample of 12 articles, there were still instances of heterogeneity in geographic location, school size, method of measuring outcome, curriculum topic among others. In this way, although the structure of the study is relatively similar, there may be confounding variables not accounted for. Furthermore, the outcome measures for specifically Group 2 studies were too specialized, in that we could not generalize them under one topic and compare them amongst Group 1 studies. Our comparison is not complete without getting enough reports under satisfaction, engagement, participation, and fidelity measures. Likewise Group 1 studies did not cover the variety of outcomes that Group 2 articles did: the Group 1 analysis was therefore more surface level in determining effectiveness of adolescent wellbeing.

In summary, across the curriculum literacy outcome measure, both Group 1 and Group 2 modes of teaching were moderate to highly effective in retention and learning of curriculum material. Whether from either approach, our findings suggest that facilitator-led mental health education programs have great potential for efficient and effective dissemination of mental health literacy. Teacher-only training sessions were well-received, demonstrating high satisfaction, engagement, fidelity, and suggesting promising evidence for effective integration at the school level. However, measured impact among adolescents still remains unclear without direct outcomes responded by the adolescents themselves. In the teacher and adolescent training programs, there were more revealing with specific outcomes better tailored to the program and response for adolescents. Some studies found clear improvements in attitudes and emotional regulations, especially among particular demographics of student populations. With this diverse array of outcomes, it was more difficult to generalize results based on similar factors, therefore making the comparison between Group 1 and Group 2 studies challenging. Nevertheless, interventions enhancing school mental health professional or general school educator mental health literacy through a Project ECHO-like model could potentially lay the groundwork for improved youth wellbeing and engagement, underscoring the importance in fostering resilience among community wide evidence-based practices. Application of these interventions demonstrated most notably improved curriculum understanding levels as compared to their control groups’ baseline scores, although the success of which may be limited to the number of studies implemented, the role of student engagement, and the mode of measuring outcomes.

Future research should aim to standardize outcome measures while also allowing for diversity, maximizing the ability to address mental health and compare research more effectively. Refining interventions to increase engagement and effectiveness will help enhance both Group 1 and Group 2 approaches to mental health education. Altogether, more research and interventions need to be conducted to better examine the relationship between subgroups of students and modes of learning models.

## Data Availability

All data produced in the present work are contained in the manuscript.

